# Interactions between SARS-CoV-2 and Influenza and the impact of coinfection on disease severity: A test negative design

**DOI:** 10.1101/2020.09.18.20189647

**Authors:** J Stowe, E Tessier, H Zhao, R Guy, B Muller-Pebody, M Zambon, N Andrews, M Ramsay, J Lopez Bernal

## Abstract

**Background:** The potential impact of COVID-19 alongside influenza on morbidity, mortality and health service capacity is a major concern as the Northern Hemisphere winter approaches. This study investigates the interaction between influenza and COVID-19 during the latter part of the 2019-20 influenza season in England.

**Methods:** Individuals tested for influenza and SARS-CoV-2 were extracted from national surveillance systems between 20/01/2020 and 25/04/2020. To estimate influenza infection on the risk of SARS-CoV-2 infection, univariable and multivariable analyses on the odds of SARS-CoV-2 in those who tested positive for influenza compared to those who tested negative for influenza. To assess whether a coinfection was associated with severe SARS-CoV-2 outcome, univariable and multivariable analyses on the odds of death adjusted for age, sex, ethnicity, comorbidity and coinfection status.

**Findings:** The risk of testing positive for SARS-CoV-2 was 58% lower among influenza positive cases, suggesting possible pathogenic competition between the two viruses. Patients with a coinfection had a risk of death of 5.92 (95% CI, 3.21-10.91) times greater than among those with neither influenza nor SARS-CoV-2 and 2.27 (95% CI, 1.23 to 4.19) greater than COVID alone suggesting possible synergistic effects in coinfected individuals. The odds of ventilator use or death and ICU admission or death was greatest among coinfection patients showing strong evidence of an interaction effect compared to SARS-CoV-2/influenza acting independently.

**Interpretation:** Cocirculation of these viruses could have a significant impact on morbidity, mortality and health service demand. Testing for influenza alongside SARS-CoV-2 and maximising influenza vaccine uptake should be prioritised to mitigate these risks.

**Funding:** This study was funded by Public Health England

## Introduction

It is likely that both SARS-CoV-2 and seasonal respiratory pathogens, most notably influenza, will be co-circulating as the northern hemisphere 2020/21 winter approaches. The potential impact of COVID-19 alongside influenza on morbidity, mortality and health service capacity is a major concern, however, currently little is understood about the interaction between these two respiratory viruses^1,2^.

There is existing evidence of pathogenic competition between respiratory viruses, including between influenza and seasonal coronaviruses^3-5^. This could be through immune-mediated interference resulting in some viruses to diminish during the peak of another virus, a phenomenon that has been recognised for many decades ^3,6,7^ One study reported that influenza vaccination was associated with an increased risk of seasonal coronavirus ^5^. To date there is some evidence of ectopic interaction between the SARS-COV-2 protein and host proteins ^8^, however there is no information on the pathogenic interaction between SARS-CoV-2 and influenza and the epidemiological impact of such interaction is unknown.

If individuals are coinfected with both SARS-CoV-2 and influenza, this could lead to more severe disease outcomes. Since the beginning of the SARS-CoV-2 pandemic, a number of case reports of SARS-CoV-2 and influenza coinfection with severe outcomes have been published ^1,9-13^ However, there is a propensity for case reports to highlight more severe cases and there has been no systematic analysis of disease outcomes in coinfected patients compared to non-coinfected controls.

In the UK the 2019-2020 influenza season peaked early with activity declining significantly from January 2020 ^14^. The season saw lower activity with influenza A(H3N2) as the predominant strain ^14^. The first SARS-CoV-2 infection occurred in late January 2020 arising from an imported case, and the distribution of SARS-CoV-2 rose with sustained community transmission from early March in the UK peaking on 7 April 2020 with 4,493 cases and on 21 April the total number of daily SARS-CoV-2 deaths peaked at 1,172 ^15^. As such there was only a limited period of overlap between influenza circulation and SARS-CoV-2 circulation. In this study, we explore the interaction between influenza and SARS-CoV-2 during the latter stages of the 2019-2020 influenza season in England.

The aims of the study are two-fold; firstly, to assess whether infection with influenza is associated with a reduced risk of SARS-CoV-2 infection and secondly to assess whether coinfection with influenza is associated with a more severe SARS-CoV-2 outcome such as death, being admitted to hospital, admitted to ICU or requiring ventilatory support.

## Methods

### Data sources and data linkage

The SGSS (Second Generation Surveillance System) and DataMart were used to obtain all influenza positive cases between 01/01/2020 and 02/06/2020 ^16,17^. For the analyses data was restricted to the time period between 20/01/2020 up to 25/04/2020, when the first SARS-CoV-2 and influenza coinfection occurred and the last influenza sample was reported in DataMart. Individuals tested for influenza who had a negative result in DataMart were also extracted. Both groups were matched to SARS-CoV-2 test results (positive and negative) in SGSS as of 02/06/2020. A coinfection was defined as positive for both influenza and SARS-CoV-2 within 7 days of each sample date.

Cases of SARS-CoV-2 and influenza coinfection were matched to the Public Health England COVID-19 deaths dataset. In addition, all cases were linked to the Demographic Batch Service (DBS), a national database coordinated by NHS digital that allows the tracing of information against personal demographics, and the date of death was extracted ^18^. Deaths from 6 days before to 28 days after the test result were included.

Test results were also linked to the Secondary Uses Service (SUS) dataset and the Hospital Episode Statistics dataset, which contain information on all admitted patient care, outpatient and A&E attendances at NHS hospitals in England ^19 20^. These datasets were used to identify patients in an Intensive Care Unit (ICU) and that required the use of a ventilator within 14 days before to 28 days after the earliest test sample date were as outcome variables as well as ethnicity^21^ and comorbidities as covariates. Comorbidities were identified using the International Classification of Diseases 10^th^ revision (ICD-10) codes and grouped into the following categories: asplenia or dysfunction of the spleen, asthma, chronic heart disease, chronic kidney disease, chronic liver disease, chronic neurological disorders, chronic respiratory disease (excluding asthma), dementia including Alzheimer’s, diabetes, malignancies affecting the immune system, obesity, other neoplasms, rheumatological diseases, and transplantations and conditions affecting the immune system. For the comorbidities linkage, data was restricted to inpatient and outpatient hospital episodes in the last 5 years.

### Statistical analysis

#### Effect of influenza infection on the risk of SARS-CoV-2 infection

The total number of positive and negative SARS-CoV-2 and influenza test results from weeks 1 to 17, 2020 were assessed. Percent positivity was calculated for individuals with a SARS-CoV-2 and influenza coinfection and individuals with no influenza infection by dividing the number of individuals with SARS-CoV-2 positive results by the total number of individuals tested and multiplied by 100. Additionally, the total number individuals with a SARS-CoV-2 and influenza coinfection were assessed by influenza type.

To estimate the effect of recent influenza infection on the risk of SARS-CoV-2 infection, univariable and multivariable analyses on the odds of SARS-CoV-2 in those who tested positive for influenza compared to those who tested negative for influenza were conducted adjusting for age, sex, ethnicity, region, comorbidity and sample week. Finally, to determine the influence of unmeasured confounding such as occupation, the analysis was stratified by age into children (under 19 years), working age adults (19-65) and older adults (>65).

#### Severity and risk of death among individuals with a coinfection

The mortality rate among individuals with a SARS-CoV-2 and influenza coinfection and those with SARS-CoV-2 infection who tested negative for influenza was calculated by dividing the number of deaths by the total number of individuals tested by age group.

To assess whether having a coinfection was associated with death, univariable and multivariable analyses on the odds of death adjusted for age, sex, ethnicity, comorbidity (0 or 1+) and coinfection status (Flu negative/ SARS-CoV-2 negative; Flu negative/ SARS-CoV-2 positive; Flu positive/ SARS-CoV-2 negative; Flu positive/ SARS-CoV-2 positive) was assessed. This analysis was repeated with a composite outcome of ventilator use or death use and a composite outcome of ICU admission or death.

## Results

A total of 19,256 individuals were tested for both influenza and SARS-CoV-2 between 20/01/2020 and 25/04/2020, when the last positive influenza test was detected in Datamart. In total, 58 individuals had a SARS-CoV-2 and influenza coinfection, 992 had a positive influenza result and were negative for SARS-CoV-2, 4,443 had a positive SARS-CoV-2 result and were negative for influenza and the remaining 13,763 were negative for both SARS-CoV-2 and influenza during this period (Figure 1). Of the 58 patients with a SARS-CoV-2 and influenza co-infection 32 (55.2%) cases were ages 70 years and older. Of the 58 cases with a SARS-CoV-2 and influenza coinfection, 31 individuals had influenza type A (unsubtyped), 16 had influenza type B, 8 had influenza H1N1, one had influenza type A&B and two cases had unknown influenza type. Week 12 had the highest reported SARS-CoV-2 and influenza coinfections (20 individuals, Table 1).

**Figure 1:**
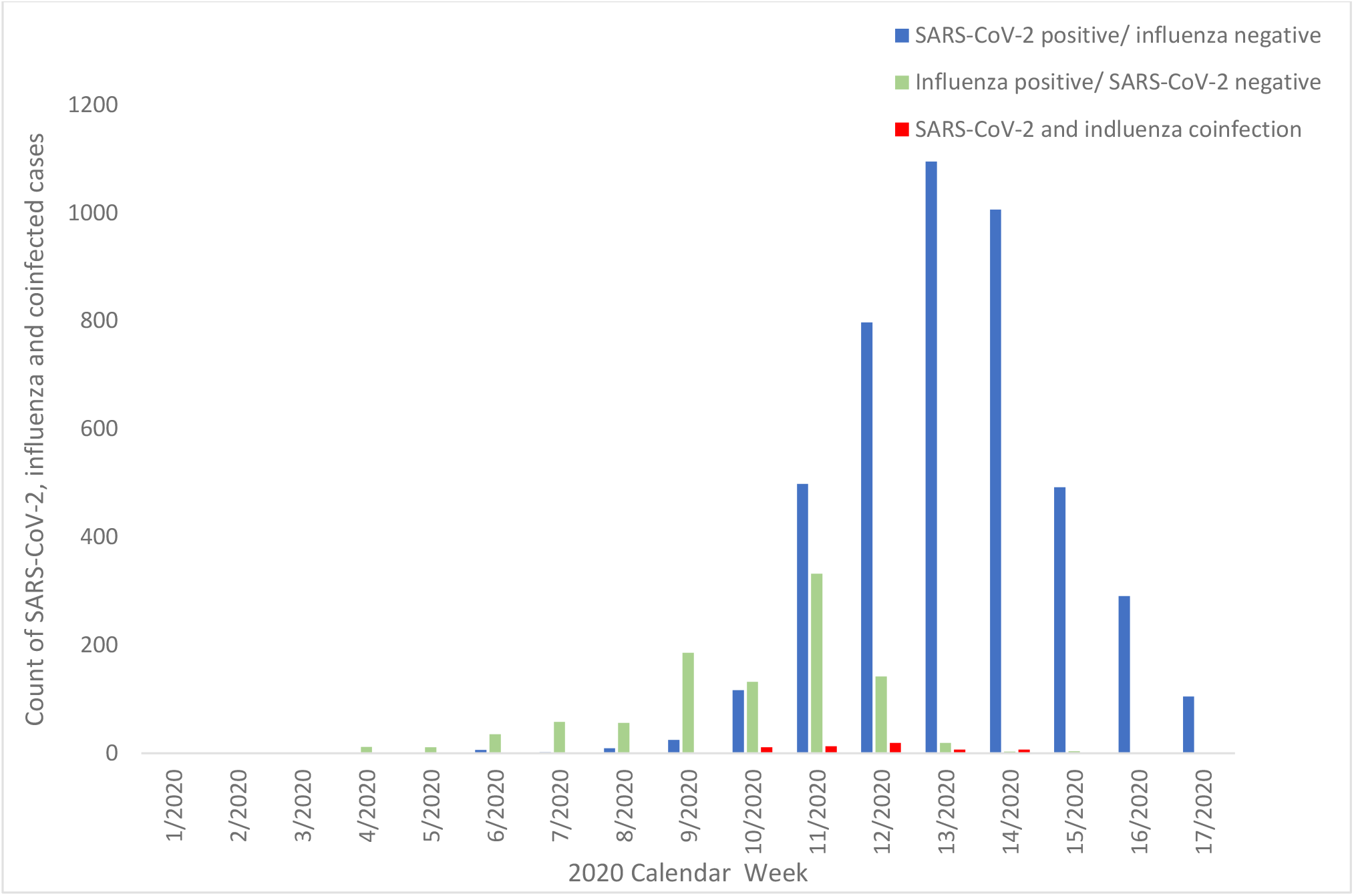
Distribution of SARS-CoV-2, influenza and coinfected cases in England from between 20/01/2020 and 25/04/2020 (Weeks 1 −17).

**Table 1.** SARS-CoV-2 positivity among influenza cases and influenza test negatives by sample week in England from 20/01/2020 to 25/04/2020

| Week | Influenza Positive |  |  |  | Influenza Negative |  |  |  |
| --- | --- | --- | --- | --- | --- | --- | --- | --- |
|  | SARS-CoV-2 - Negative | SARS-CoV-2 - Positive | Total | SARS-CoV-2 Positivity (%) | SARS-CoV-2 - Negative | SARS-CoV-2 - Positive | Total | SARS-CoV-2 Positivity (%) |
| 1 | 1 | 0 | 1 | 0 | 0 | 0 | 0 | - |
| 2 | 0 | 0 | 0 | - | 1 | 0 | 1 | 0 |
| 3 | 0 | 0 | 0 | - | 4 | 0 | 4 | 0 |
| 4 | 12 | 0 | 12 | 0 | 34 | 0 | 34 | 0 |
| 5 | 11 | 0 | 11 | 0 | 60 | 0 | 60 | 0 |
| 6 | 34 | 0 | 34 | 0 | 323 | 6 | 329 | 1.8 |
| 7 | 58 | 0 | 58 | 0 | 583 | 2 | 585 | 0.3 |
| 8 | 53 | 0 | 53 | 0 | 329 | 9 | 338 | 2.7 |
| 9 | 189 | 0 | 189 | 0 | 1471 | 22 | 1493 | 1.5 |
| 10 | 125 | 11 | 136 | 8.1 | 1211 | 113 | 1324 | 8.5 |
| 11 | 330 | 12 | 342 | 3.5 | 2957 | 483 | 3440 | 14.0 |
| 12 | 145 | 20 | 165 | 12.1 | 2322 | 783 | 3105 | 25.2 |
| 13 | 25 | 6 | 31 | 19.4 | 1342 | 1092 | 2434 | 44.9 |
| 14 | 4 | 8 | 12 | 66.7 | 1072 | 1022 | 2094 | 48.8 |
| 15 | 4 | 1 | 5 | 20.0 | 839 | 496 | 1335 | 37.2 |
| 16 | 0 | 0 | 0 | - | 758 | 303 | 1061 | 28.6 |
| 17 | 1 | 0 | 1 | 0 | 451 | 111 | 562 | 19.8 |

A total of 13,451 (70%) individuals linked to a hospital admission record in SUS between 01/12/2020 and 24/08/2020 of which 12,253 individuals had an associated record in the 14 days before and up to 28 days after the earliest SARS-CoV-2 or influenza test date. A total 1,666 (6%) of individuals had an ICU admission and 890 (5%) were ventilated (Supplementary Figure 1). Of the 19,256 cases, 2,469 (12.8%) died of which 25/58 (43.1%) of the SARS-CoV-2 and influenza coinfected cases died.

### Effect of influenza infection on the risk of SARS-CoV-2 infection

SARS-CoV-2 positivity among influenza positive cases was generally lower than SARS-CoV-2 positivity among influenza test negatives (Table 1). The highest SARS-CoV-2 positivity rate for both influenza positive and negative cases was in week 14 (66.7% and 44.8%, respectively).

After adjusting for age, sex, ethnicity, region, comorbidity and sample week in the multivariable analysis, the results indicate that the odds of testing positive for SARS-CoV-2 was 58% lower among influenza positive cases (OR 0.42, 95% CI 0.31-0.56) (Table 2). After stratifying by age into children (under 19 years), working age adults (19-65) and older adults (>65), the working age and older population had a significantly lower odds of SARS-CoV-2 if testing positive for influenza (OR 0.26 (95% CI 0.15-0.45) and (OR 0.52 (95% CI 0.35-0.75)), respectively. Conversely, there was no association between influenza positivity and SARS-CoV-2 positivity among children (OR 1.07 (95% CI 0.38 – 3.01) though numbers were small in this age group. To formally test the interaction between influenza and the stratified age cohorts, the model was fitted with separate terms for the age cohorts resulting in no evidence of interaction (p=0.01).

**Table 2.**
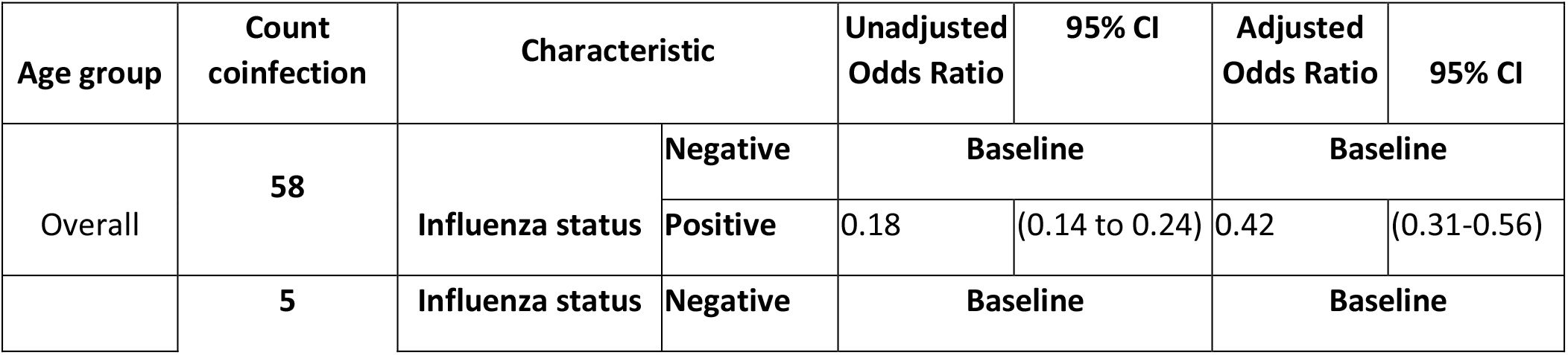
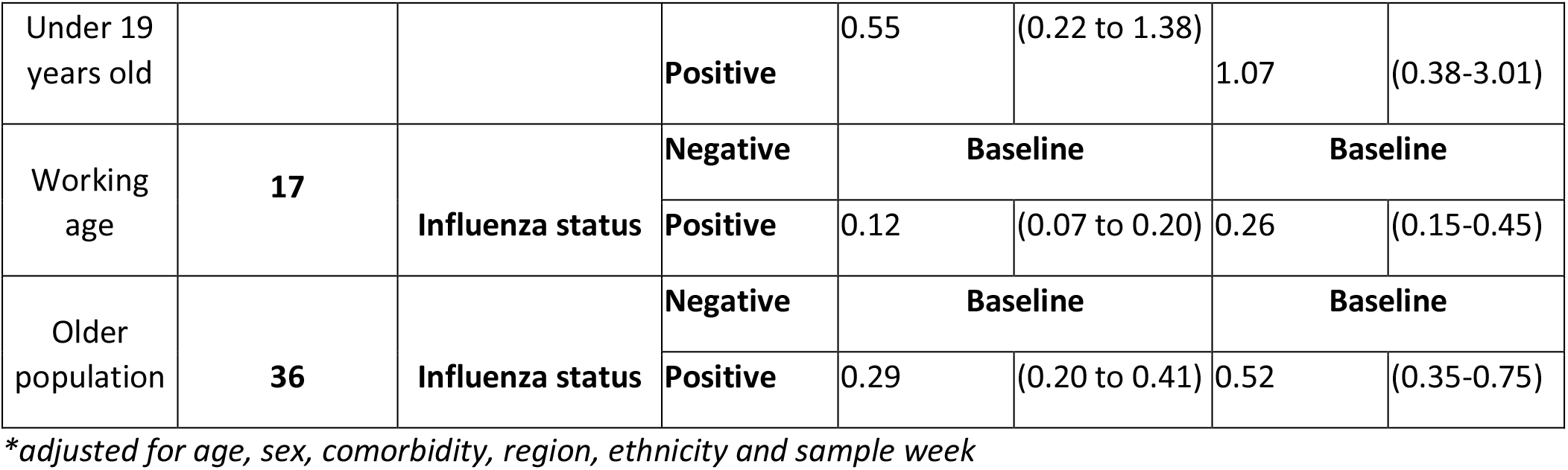
Odds of SARS-CoV-2 infection by influenza status stratified by age (England from 20/01/2020 to 25/04/2020)*

### Risk of death among individuals with a coinfection

After linking all individuals to the death datasets, a total of 2,699 individuals had a recorded death with a SARS-CoV-2 or influenza test (positive or negative) within 28 days before and six days after the death date. Of the reported deaths, 26 (1.0%) individuals had a SARS-CoV-2 and influenza coinfection, 1,419 (52.6%) had a SARS-CoV-2 infection only, 48 (1.8%) had influenza only and 1,206 (44.7%) had neither a SARS-CoV-2 or influenza positive results.

Overall 43.1% of cases with coinfection died compared to 26.9% of those who tested positive only for SARS-CoV-2 (Table 3). Age specific mortality rates were higher among older people with a SARS-CoV-2 and influenza coinfection (Table 3). For individuals with influenza only, the overall mortality rate was 48/992=4.8% and for those negative for both, the mortality rate was 1,203/13,763=8.7%.

**Table 3.** SARS-CoV-2 and influenza coinfection deaths and mortality rate (%) and COVID-19 with no influenza deaths and mortality rate (%) by age groups in England from 20/01/2020 to 25/04/2020

| Age | Coinfection (Flu positive and SARS-CoV-2 positive) n=58 |  |  |  |  | Single infection (SARS-CoV-2 positive and Flu negative) |  |  |  |  |
| --- | --- | --- | --- | --- | --- | --- | --- | --- | --- | --- |
|  | Total | Died | ICU admission | Mortality Rate (%) | ICU Rate (%) | Total | Died | ICU admission | Mortality Rate (%) | ICU Rate (%) |
| <5 | 0 | 0 | 0 | - | - | 37 | 0 | 2 | 0.0 | 5.4 |
| 5-9y | 2 | 0 | 0 | 0.0 | 0.0 | 7 | 0 | 0 | 0.0 | 0.0 |
| 10-19y | 3 | 0 | 0 | 0.0 | 0.0 | 33 | 1 | 4 | 3.0 | 12.1 |
| 20-29y | 1 | 0 | 0 | 0.0 | 0.0 | 162 | 4 | 10 | 2.5 | 6.2 |
| 30-39y | 7 | 1 | 2 | 14.3 | 28.6 | 295 | 5 | 33 | 1.7 | 11.2 |
| 40-49y | 2 | 0 | 0 | 0.0 | 0.0 | 426 | 21 | 80 | 4.9 | 18.8 |
| 50-59y | 5 | 1 | 3 | 20.0 | 60.0 | 658 | 81 | 158 | 12.3 | 24.0 |
| 60-69y | 6 | 3 | 1 | 50.0 | 16.7 | 670 | 155 | 160 | 23.1 | 23.9 |
| 70-79y | 14 | 8 | 1 | 57.1 | 7.1 | 824 | 307 | 117 | 37.3 | 14.2 |
| 80+ | 18 | 12 | 0 | 66.7 | 0.0 | 1,331 | 619 | 15 | 46.5 | 1.1 |
| **Total** | **58** | **25** | **7** | **43.1** | **12.1** | **4,443** | **1,193** | **581** | **26.9** | **13.1** |

The multivariable analysis adjusting for age, sex, ethnicity, comorbidity (0 or 1+) and coinfection status (Flu negative/ SARS-CoV-2 negative; Flu negative/ SARS-CoV-2; Flu positive/ SARS-CoV-2 negative; Flu positive/ SARS-CoV-2 positive) indicated that

The odds of death was 5.92 times greater among individuals with a SARS-CoV-2 and influenza coinfection than those with neither influenza nor SARS-CoV-2 (95% CI 3.21-10.91) and was higher than those with only COVID-19 where the odds of death was 2.61 time greater compared to no SARS-CoV-2 or influenza (Table 4). For those only positive for influenza there was a slightly decreased mortality risk (OR 0.64 (95% CI 0.47-0.89)). Furthermore, patients with a SARS-CoV-2 and influenza coinfection were around 2.3 times (95% CI, 1.23 to 4.19) more likely to die compared to those with SARS-CoV-2 alone. To formally test the interaction between influenza and SARS-CoV-2 the same model was fitted but with separate terms for influenza, SARS-CoV-2 and the interaction of influenza and SARS-CoV-2, this gave a significant interaction effect (P=<0.001) of an additional 3.60 odds of death (95% CI 1.83-7.11) compared to that expected if influenza and SARS-CoV-2 acted independently.

**Table 4:**
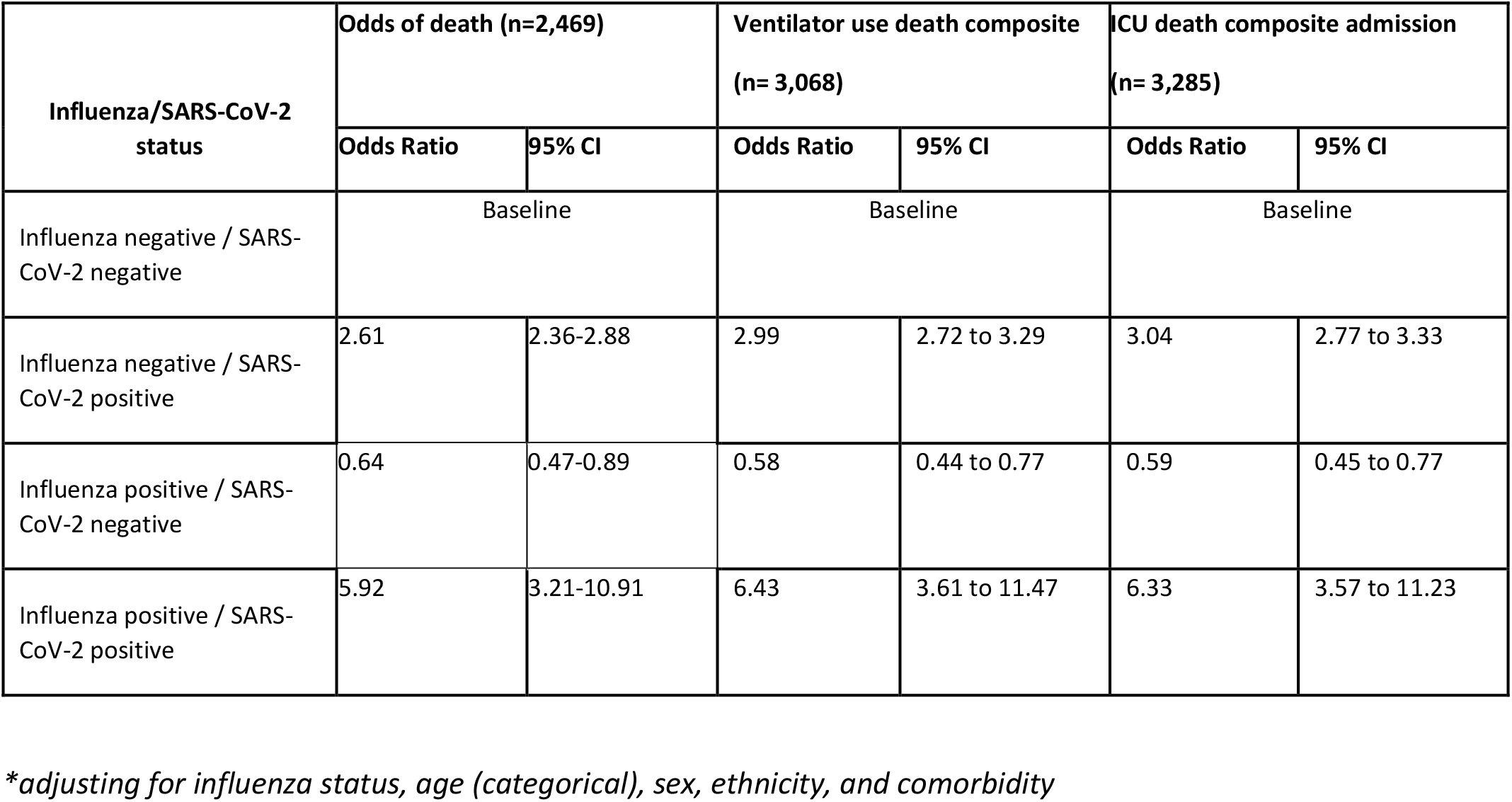
Odds of hospital admission, ICU admission ventilator use and death by influenza/SARS-CoV-2 status in England from 20/01/2020 to 25/04/2020*

When combining ventilator use or death into a composite variable, the odds was 6.43 times greater among individuals with coinfection (95% CI 3.61-11.47). The ICU admission or death composite had an odds 6.33 times greater among individuals with coinfection (95% CI 3.57-11.23) (Table 4). A test for interaction for both the ventilator composite and ICU composite gave a significant effect (P=<0.001) with additional 3.38 odds of coinfection (95% CI 1.81-6.34) and 3.39 odds of coinfection(95% CI 1.83-6.29), respectively compared to that expected if influenza and SARS-CoV-2 acted independently.

## Discussion

We found that influenza infection was associated with a lower risk of SARS-CoV-2 infection, suggesting that there may be pathogenic competition between these two viruses. We also found strong evidence that coinfection with influenza and SARS-CoV-2 was associated with an increased risk of death or severe disease and that this appears to be beyond the additive effect of the two viruses acting independently.

The risk of testing positive for SARS-CoV-2 was 58% lower among influenza positive cases. This is consistent with recent descriptive evidence from New York where <3% of those testing positive for SARS-CoV-2 had coinfection with influenza whereas 13% of those testing negative for SARS-CoV-2 were influenza positive ^22^. It is also consistent with existing evidence on the interaction between influenza and seasonal coronavirus and rhinovirus ^3-5,23^. There are biologically plausible mechanisms for such an effect, including stimulation of non-specific immune responses by the first infectious agent, such as the induction of a refractory state in bystander cells as a result of the antiviral effect of interferon induced as part of an innate immune response to an RNA viral infection.

Our findings cannot distinguish between a reduced risk of SARS-CoV-2 among those first infected with influenza or vice versa. A recent study has suggests that SARS-CoV-2 has a lower growth rate than influenza and is suppressed if the infections start simultaneously, however, if an influenza infection were to occur after SARS-CoV-2 infection, a coinfection would be detected ^24^. Our findings would not support the relaxation of preventative measures against influenza, including vaccination, given the risk of morbidity and mortality from influenza ^5,25,26^ as well as our finding of adverse outcomes associated with influenza and SARS-CoV-2 coinfection. Furthermore, results from Brazil indicated a significantly lower odds of needing intensive care treatment, invasive respiratory support and death among patients with SARS-CoV-2 that received the inactivated trivalent influenza vaccine 27. The International Council on Adult Immunization highlights in their roadmap that influenza, pneumococcal and herpes zoster vaccines programmes are more urgent than ever before ^28^. As a further potential implication on influenza vaccination, if there is a competitive effect between influenza and SARS-CoV-2, this effect may also be seen with live attenuated influenza vaccination (LAIV) which if offered to children in England and could in turn have a role in outbreak management. Further research on the pathology of influenza and SARS-CoV-2 coinfection such as the order of infection and the effect of timing of influenza infection on the risk of acquiring SARS-CoV-2 infection, as well as any effect of LAIV is required.

The results from this study indicate that the risk of death was nearly six times greater among individuals with a SARS-CoV-2 and influenza coinfection than those with neither influenza nor SARS-CoV-2 and that this effect is significantly higher than the risk associated with SARS-CoV-2 infection alone. Similarly, the combined outcomes of ventilator use or death and ICU admission or death gave similar results. These findings suggest a possible synergistic effect between SARS-CoV-2 and influenza once an individual is coinfected. The high mortality rate is consistent with case reports of severe outcomes in coinfected patients ^12,13,29^. Conversely, some case series have not seen increased severity with influenza and SARS-CoV-2 co-infection, where the outcomes have been similar to cases with SARS-CoV-2 only ^30,31^. Synergistic effects have previously been reported between influenza and other respiratory viruses, for example by facilitating cell to cell spread ^32^. These findings also emphasise the importance of influenza vaccination in at risk groups and early administration of antivirals where coinfection is identified or suspected. This also adds further weight to the need for effective vaccines against influenza, in particular among the elderly among whom vaccine effectiveness tends to be lower and among whom most coinfections were seen. This has been an area of development in recent years with the introduction of high dose and adjuvanted vaccines^33^.

Studies of other respiratory viral infections have not indicated adverse outcomes from coinfection, for example, a study assessing SARS-CoV and metapneumovirus in Hong Kong that showed that there was no significant difference in the outcomes, including deaths between those with a SARS-CoV and metapneumovirus coinfection versus SARS-CoV alone ^34^. It is important to note, that these are case studies of hospitalised individuals and the comparisons do not adjust for potential confounders.

To our knowledge, our study is the first epidemiological study that uses national level data on both positive and negative SARS-CoV-2 and influenza cases. By extracting all cases with a SARS-CoV-2 and influenza test result, and linking the data to HES we were able to assess the effects of SARS-CoV-2 and influenza co-infections compared to single infection and negative test results while controlling for variables such as ethnicity, comorbidities, sex and age which are known factors for SARS-CoV-2 morbidity ^35-37^. Furthermore, the test negative design controls for the propensity for more severe cases to be tested for other respiratory viruses.

Most of the SARS-CoV-2 tests were collected when the government policy was to test individuals on admission to hospital with lower respiratory tract infections and healthcare workers ^38^. Therefore, the majority of SARS-CoV-2 cases were individuals with moderate to severe symptoms and mild cases are likely to be missed. Additionally, influenza test results collected from DataMart are only collected from sentinel laboratories. However, the test negative controlled design means that none of the study arms were biased towards more severe outcomes as all were tested for both diseases.

Additionally, in our study the majority of cases with a SARS-CoV-2 coinfection had influenza subtype A. Due to small numbers it was not possible to determine whether the risk of SARS-CoV-2 coinfection and severity of disease varied by influenza subtype. While in the 2019-2020 influenza season, the majority of influenza subtype A cases were H3N2, towards the end of the season there was a shift towards H1N1, which is consistent with our finding of more H1N1 cases among those coinfections that were subtyped ^14^. The impact severity of influenza and SARS-CoV-2 coinfection by different subtypes should be further considered in the upcoming influenza season. Furthermore, the influenza vaccination status of the patients was not available therefore we could not adjust for vaccination status of the patients in the model. While our findings provide evidence of pathogenic competition between influenza and SARS-CoV-2, a significant number of coinfections occur and they appear to be associated with higher mortality rates. Further investigation is needed in order to understand the potential mechanisms for any synergistic interaction.

Cocirculation of these two viruses could have a significant impact on morbidity, mortality and health service demand. As the 2020-2021 northern hemisphere influenza season approaches, it is important that a high index of suspicion for coinfection is maintained. Testing strategies should include influenza and other respiratory viruses as well as SARS-CoV-2 and measures should be adopted to prevent coinfection including maximising uptake of influenza vaccination, particularly in groups at higher risk of both diseases.

## Data Availability

The SGSS (Second Generation Surveillance System) and DataMart were used to obtain all influenza positive cases. Cases of SARS-CoV-2 and influenza coinfection were matched to the Public Health England COVID-19 deaths dataset. In addition, all cases were linked to the Demographic Batch Service (DBS), a national database coordinated by NHS digital that allows the tracing of information against personal demographics, and the date of death was extracted.
Test results were also linked to the Secondary Uses Service (SUS) dataset and the Hospital Episode Statistics dataset, which contain information on all admitted patient care, outpatient and A&E attendances at NHS hospitals in England.
All PHE individual-level owned databases used in this study were under Regulation 3 of The Health Service (Control of Patient Information) (Secretary of State for Health, 2002). Availability of this data could lead to identifiable deductive disclosure and therefore is not shared in the public domain. 
For all other datasets used in this study, links for user permissions can be found here:
DBS: http://nww.connectingforhealth.nhs.uk/demographics/dbs/.
HES: https://digital.nhs.uk/data-and-information/data-tools-and-services/data-services/hospital-episode-statistics.
SUS: https://digital.nhs.uk/services/secondary-uses-service-sus

http://nww.connectingforhealth.nhs.uk/demographics/dbs/

## Author contributions

Authors JLB, ET, JS, NA developed the study protocol.

Authors HZ, RG, JS, ET, BMP extracted the data.

Authors JS, ET, JLB, NA conducted the statistical analysis

Authors ET, JS, NA, JLB, MR, BMP, RG, HZ, MZ data interpretation,

Authors ET, JS, NA, JLB, MR, BMP, HZ wrote the first draft of the paper.

All authors contributed to subsequent revisions and approved the final version.

